# A rare coding mutation in the MAST2 gene causes venous thrombosis in a French family with unexplained thrombophilia: The Breizh MAST2 Arg89Gln variant

**DOI:** 10.1101/2020.05.23.20101402

**Authors:** P.-E. Morange, F. Peiretti, L. Gourhant, C. Proust, A-S Pulcrano-Nicolas, G.-V. Saripella, L. Stefanucci, R. Lacroix, M. -Ibrahim.Kosta, C. Lemarié, Mattia Frontini, M.-C. Alessi, D.-A. Trégouët, F. Couturaud

## Abstract

Rare variants outside the classical coagulation cascade might cause rare inherited thrombosis. We aimed to identify the variant(s) causing venous thromboembolism (VTE) in a family with multiple relatives affected with unprovoked VTE and no thrombophilia defects. We identified by whole exome sequencing an extremely rare Arg to Gln variant (R89Q) in the Microtubule Associated Serine/Threonine Kinase 2 *(MAST2)* gene that segregates with VTE in the family. Free-tissue factor pathway inhibitor (f-TFPI) plasma levels were significantly decreased in affected family members compared to healthy relatives. Conversely, plasminogen activator inhibitor-1 (PAI-1) levels were significantly higher in affected members than in healthy relatives. RNA sequencing analysis of RNA interference experimental data conducted in endothelial cells revealed that, of the 13,387 detected expressed genes, 2,354 have their level of expression modified by MAST2 knockdown, including *SERPINE1* coding for PAI-1 and *TFPI*. In HEK293 cells overexpressing the MAST2R289Q variant, *TFPI* and *SERPINE1* promoter activities were respectively lower and higher than in cells overexpressing the MAST2 wild type allele. This study identifies a novel thrombophilia-causing R89Q variant in the *MAST2* gene that is here proposed as a new molecular player in the etiology of VTE by interfering with hemostatic balance of endothelial cells.

**Author Summary:** Venous thromboembolism (VTE) is a multifactorial disease in which the genetic burden. We here present the case of a French family with multiple relatives affected with unprovoked VTE (i.e. that occurred in the absence of clinical risk factors) in which no thrombophilia defects had been identified. Adopting a whole exome sequencing approach, we identified an extremely rare variant located in the Microtubule-associated serine/threonine-protein kinase-2 *(MAST2)* gene that perfectly segregates with the VTE phenotype and that interferes with hemostatic balance of endothelial cells. Our results pave the way for adding *MAST2* to the list of genes to be sequenced and looked for in thrombophilia families with unprovoked VTE.

## Introduction

Venous thromboembolism (VTE) is a multifactorial disease in which the genetic burden can be characterized by a sibling relative risk of ~2.5 (1) and an estimated heritability between 35%-60% (2). As for many multifactorial diseases, the spectrum of genetic factors contributing to VTE susceptibility ranges from common single nucleotide polymorphisms (SNPs) associated with low-to-moderate genetic effects to private variants segregating within families and associated with very high relative risk of disease. About thirty common SNPs with minor allele frequency (MAF) greater than ~1% have so far been reported to associate with the risk of VTE in the general population, each of them being characterized by an Odds Ratio for disease ranging between 1.06 and 3.0 (3). Uncommon genetic variants with MAF between 0.1% and 1% have also been reported such as the *THBD* c.-151G>T variant (4) or protein S Heerlen (5). At the extreme low frequency side of the genetic spectrum reside private variants (frequencies < 1‰) that are generally ‘loss of function’ variants associated with at least a 10-fold increased risk in heterozygote individuals and mainly affecting the coagulation cascade through inherited deficiencies of the three main natural anticoagulants, antithrombin, protein C and protein S. However, rare variants outside the classical coagulation cascade have also been proposed to cause severe rare inherited thrombosis generally referred to as inherited thrombophilia (6) the identification of which being facilitated by the development of whole exome/genome sequencing technologies.

We here present the case of a French family with multiple relatives affected with unprovoked VTE (i.e. VTE that occurred in the absence of clinical risk factors) in which no thrombophilia defects had been identified. Adopting a whole exome sequencing (WES) approach, we identified an extremely rare variant located in the Microtubule-associated serine/threonine-protein kinase-2 *(MAST2)* gene that perfectly segregates with the VTE phenotype and that participates to the regulation of tissue factor pathway inhibitor (TFPI) and plasminogen activator inhibitor-1 (PAI-1) blood levels. In addition, RNA sequencing (RNA-seq) performed from endothelial cells treated with siRNAs targeting MAST2 and gene reporter experiments demonstrate a MAST2-dependent regulation of TFPI and PAI-1 gene expressions.

## Results

### Pattern of coagulation parameters in the studied family

The genealogical tree of the family with unknown thrombophilia is reported in Fig 1. Measurement of coagulation parameters in family members are reported in Table 1. While differences in prothrombin time, INR, activated partial thromboplastin time, FII, and FX were due to the use of antivitamin K in cases, unexpected nominal statistical differences (p < 0.05) were observed for plasma levels of the anticoagulant protein f-TFPI and the antifibrinolytic protein PAI-1. Plasma levels of f-TFPI were significantly (p = 0.01) lower in the affected family members compared to healthy relatives (6.6 +/-1.9 ng/mL vs 17.4 +/-1.2 ng/mL) (Fig 2) while the opposite pattern was observed for PAI-1 plasma levels (21.7 +/-6.1 IU/mL vs 3.3 +/-0.7 IU/mL, p = 0.01) (Fig 2).

**Figure 1.**
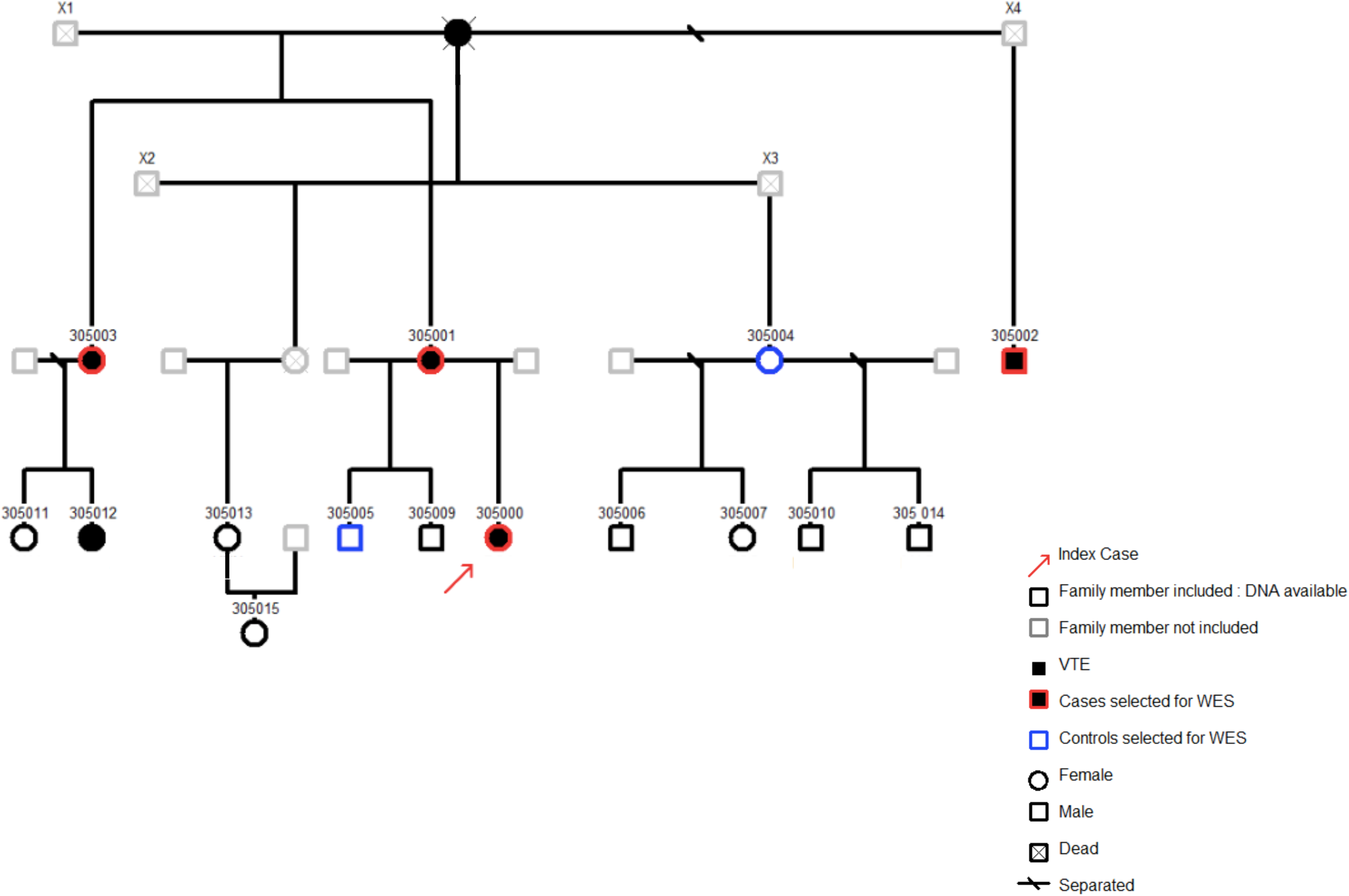
Genealogical tree of the family

**Table 1.** Measurement of coagulation parameters in the 10 subjects with citrated plasma available.

| Subjects | Status | PT (%) | INR (ratio) | aPTT (sec) | Fibrinogen (g/L) | FII (%) | FV (%) | FX (%) | f-TFPI (ng/mL) | PAI-1 (UI/mL) |
| --- | --- | --- | --- | --- | --- | --- | --- | --- | --- | --- |
| 305000 | case | 30 | 2.55 | 49.5 | 4.16 | 27 | 64 | 13 | 7.4 | 15 |
| 305001 | case | 26 | 2.88 | 46.7 | 4.3 | 18 | 76 | 9 | 2.9 | 16 |
| 305002 | case | 33 | 2.29 | 40.1 | 3.87 | 29 | 96 | 14 | 9.5 | 34 |
| 305004 | control | 98 | 1.01 | 32.4 | 4.93 | 99 | 86 | 105 | 17.2 | 1 |
| 305006 | control | 93 | 1.04 | 39 | 1.97 | 82 | 91 | 96 | 17.7 | 4 |
| 350007 | control | 100 | 1 | 35.4 | 2.62 | 107 | 75 | 122 | 15.1 | 1 |
| 305010 | control | 114 | 0.93 | 34.3 | 3.17 | 103 | 117 | 102 | 17.1 | 2 |
| 305013 | control | 104 | 0.98 | 35.1 | 3.13 | 97 | 95 | 90 | 21.9 | 4 |
| 305014 | control | 98 | 1.01 | 33.9 | 2.66 | 102 | 102 | 98 | 20.7 | 5 |
| 305015 | control | 106 | 0.97 | 37.8 | 3.26 | 95 | 91 | 95 | 12.3 | 6 |

**Figure 2.**
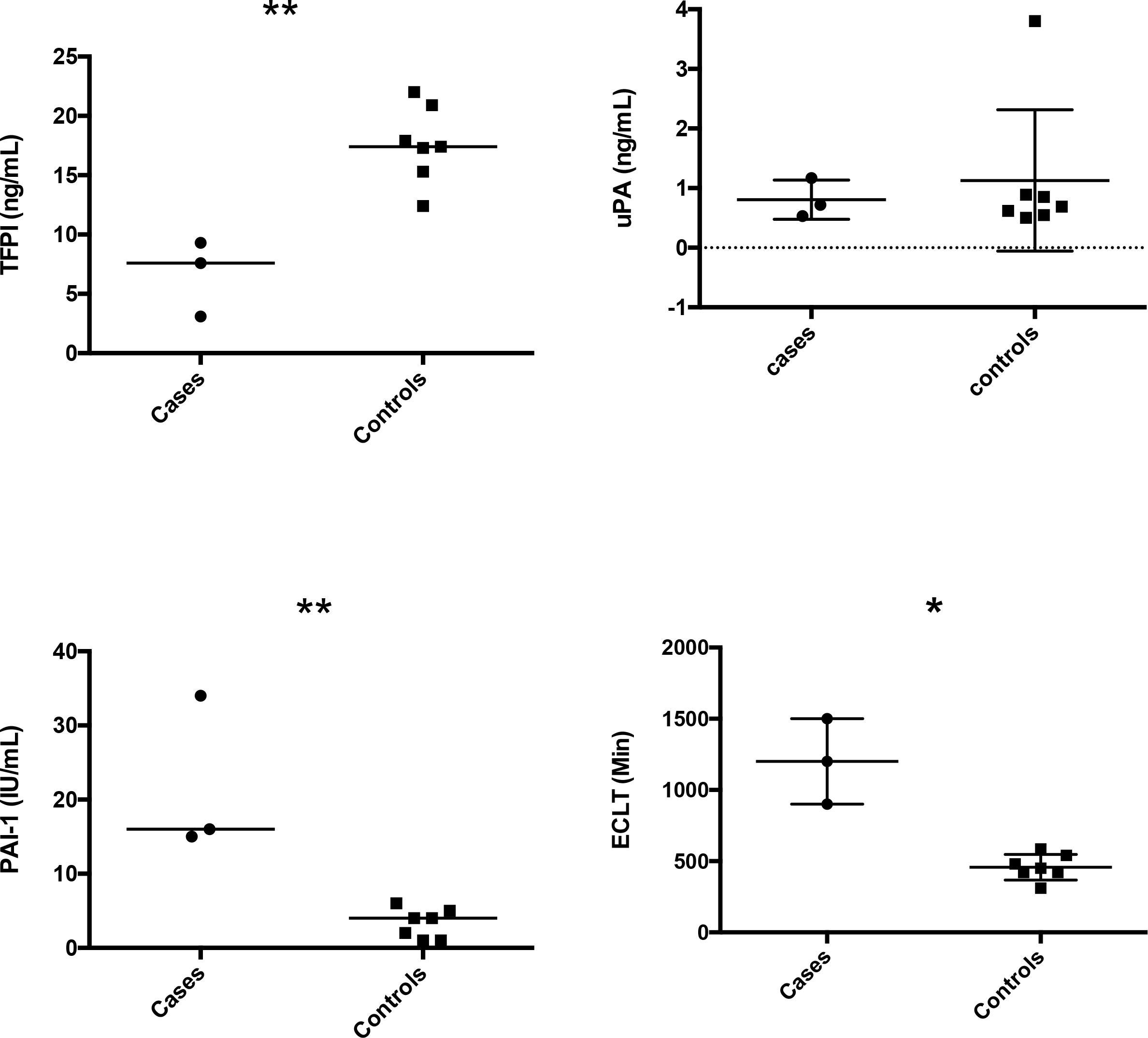
Free TFPI, euglobulin clot lysis time, PAI-1 and Urokinase plasma levels in VTE cases and controls from the family. Statistical analyses were made using Mann-Withney. * p<0.05 and **p<0.01: significant vs controls.

### Identification of a MAST2 variant following the WES strategy

As the familial transmission of the disease was compatible with an autosomal dominant inheritance, we first selected variants (n = 14,406) shared by all cases and then excluded those that were carried by controls, leaving 1,153 variants as potential candidates. Among those, 108 were kept as there were likely functional (stop loss/stop gain, frameshift insertion/deletion, non-synonymous and splicing variants). Then bioinformatics search in public genomic data repositories identified 2 as very rare candidates. One, rs151257275, was a nonsynonymous R169Q variant in the *ADAMTS10* gene, with allele frequency ~0.005 and predicted to have no deleterious effect on the protein function. The second, rs13 87081220 is a nonsynonymous Arg to Gln mutation (R89Q) in exon 2 of the *MAST2* gene and it has 2 independent heterozygous cases reported in gnomAD(7). *In silico* prediction tools PolyPhen-2, SIFT and UMD-predictor identified this mutation as deleterious. Its CADD Phred Score was 34.

These two candidate variants were then genotyped in all individuals, including those used for WES and their relatives with available DNA that were not part of the WES analysis (1 case and 8 controls). Only the *MAST2* variant perfectly co-segregates with the VTE phenotype whereas one case (305012) did not carry the *ADAMTS10* variant indicating that this variant should no longer be a candidate (S1 Table).

We genotyped this uncharacterized mutation in 6,790 VTE cases and 5,970 healthy individuals and did not detect additional carriers providing strong argument that this mutation is extremely rare. The look-up performed in other large cohort studies (i.e. 100,000 Genomes Project, gnomAD, NIHR BioResource-Rare Diseases, H3-Africa and GenomeAsia 100K; S2 Table, which includes relevant references) showed that the MAST2 Arg89Gln variant was present just in 2 alleles out of the the ~ 345,000 total exomes in these projects (S2 Table).

### Functional characterization of the mutation and its structural gene

The observation that plasma concentrations of f-TFPI and PAI-1 (two proteins produced by the endothelium) were significantly associated with the presence of the MAST2R89Q variant, led us to propose the involvement of MAST2 in regulating some endothelial properties. To gain a deeper understanding of the impact of this critical, yet relatively unknown contributor of endothelial homeostasis, an RNA-Seq based analysis was performed to compare the mRNA expression profiles of ECV 304 endothelial cells, with those of *MAST2* knockdown cells. The heatmap (Fig 3A) and volcano plot (Fig 3B) showed global expression changes in MAST2 knockdown cells. The full list of differential association p-value is given in S3 Table. Of the 13,387 expressed genes that have been detected, 2,354 have their expression modified (FDR < 1%) by *MAST2* knockdown. As expected, *MAST2* was one of the genes whose expression was the most significantly reduced (ranked 8 in terms of differential association p-value; log2 fold change (log2FC) ~ −2). The expression of *RHBLD2* was the most reduced (ranked 689; log2FC − 4.3) and that of *HIST1H2BB* the most increased (ranked 897; log2FC 3.53) by *MAST2* knockdown. The expression of some genes known to regulate the hemostatic properties of endothelial cells was altered by *MAST2* knockdown. This was the case for *SERPINE1* (coding for PAI-1) (ranked 42; log2FC —126), *PLAU* (coding for urokinase) (ranked 85; log2FC −2), *SERPINE B8* (ranked 22; log2FC 1.42) (a serine protease inhibitor reported to coexpress with *PLAU)* (https://string-db.org/cgi/input.pl), and *TFPI*(ranked 1697; log2FC ~0.60). While the expression of *PLAT* (coding for t-PA) and *THBD* (coding for thrombomodulin), two genes also involved in the regulation of endothelial function, was not significantly altered.

**Figure 3.**
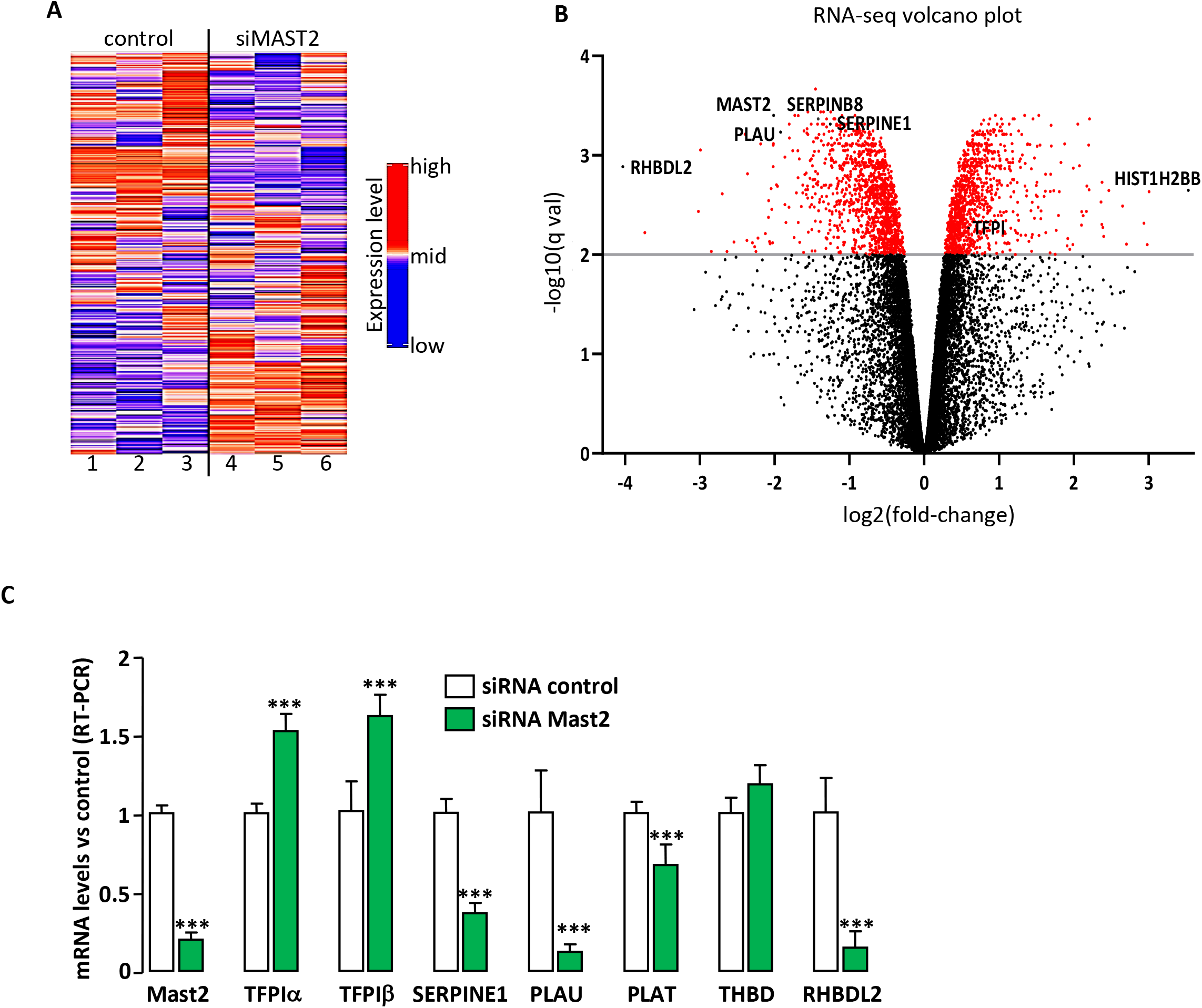
(a) Heatmap of the 5000 genes most differently expressed between cells treated with control siRNA (lanes 1-3) and with MAST2 specific siRNA (lanes 4-6). (b) volcano plot. (c) Q-PCR validation of the RNA-seq. ECV304 cells were transfected with control siRNA or with two MAST2 specific siRNA. 48 hours post transfection, mRNA levels of MAST2, TFPIα and β, SERPINE1, PLAU, PLAT, THBD and RHBLD2 were analyzed by RT-PCR. Data are means ± Standard Deviation. Statistical analyses were made using unpaired t-test. ***p<0.001: significant vs control.

Real time PCR analysis (Fig 3C) confirmed the regulation of gene expression detected by RNA-Seq, i.e. treatment of endothelial cell with siRNA specific for *MAST2* decreased the expression of *MAST2, RHBDL2, SERPINE1, PLAU* and increased the expression of TFPIα and TFPIβ. Expression of *PLAT* and *THBD* were poorly altered (Fig 3C). These results underline the ability of *MAST2* to regulate mRNA levels of genes encoding proteins involved in coagulation/fibrinolysis cascades.

Pathway analysis was then applied to the top 100 most significantly differentiated mRNA expressions according to *MAST2* silencing to assess whether this genes list could be enriched for genes belonging to specific biological pathways. Enrichment analysis was performed using the DAVID software (8) interrogating the GO, KEGG, REACTOME, and PANTHER databases. At a FDR of 1%, very few pathways were identified (Fig 4A) and S4 Table). Nevertheless, one of these pathways was significantly enriched (FDR = 0.002) for genes belonging to the fibrinolysis cascade notably *SERPINE1, PLAU* and *SERPINB8*.

**Figure 4.**
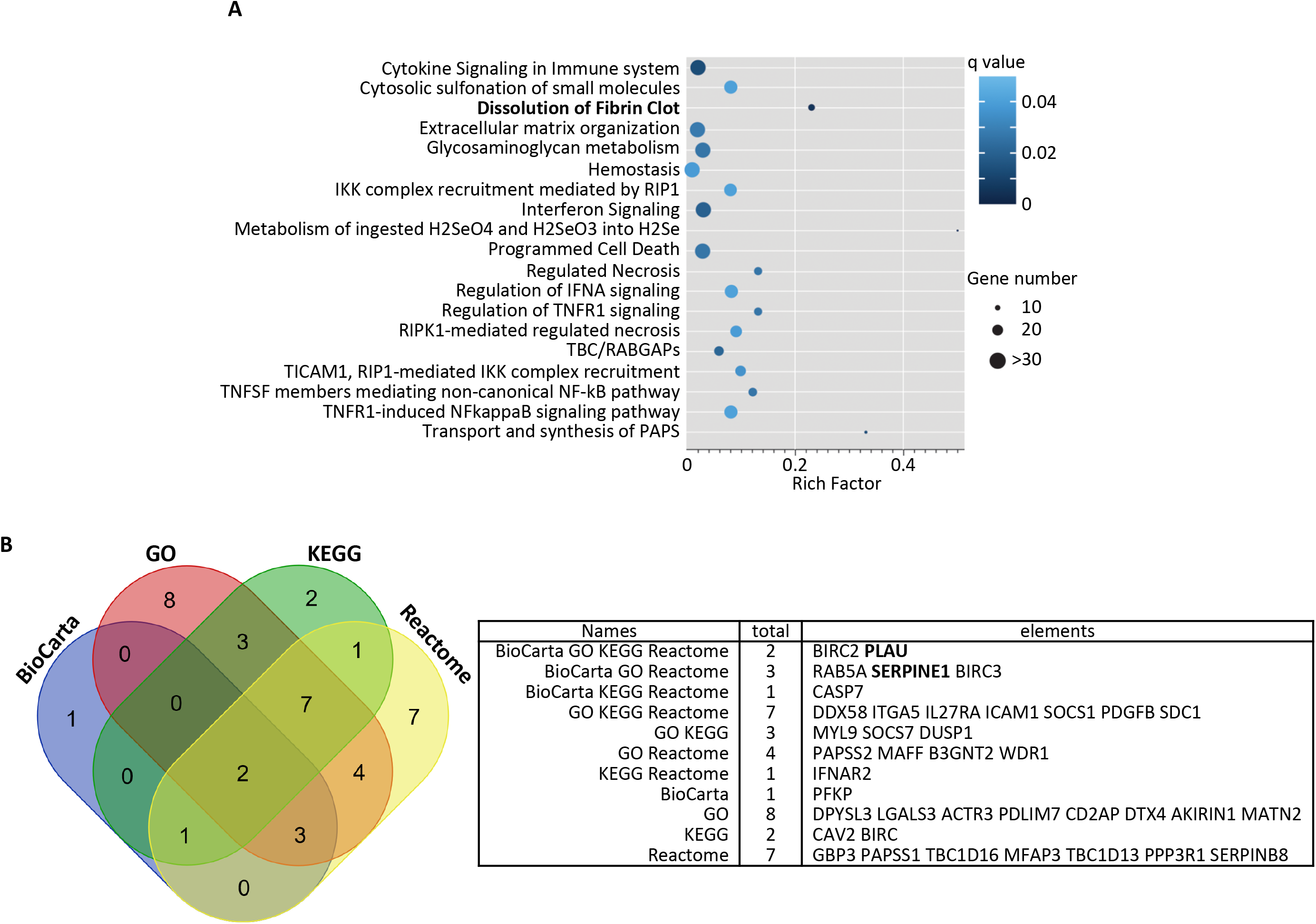
(a) Scatter plot of Reactome pathways enrichment statistics. Rich factor is the ratio of the differentially expressed gene number to the total gene number in a certain pathway. q value is corrected p value. The color and size of the dots represent the range of the q value and the number of differently expressed genes mapped to the indicated pathways, respectively. (b) Venn diagram showing the overlap of genes of the enriched biological pathways identified using BioCarta, GO, KEGG and Reactome pathway databases (supplementary Table 3).

Because pathway structure and terminology vary across databases, we determined which genes were significantly contributing to the overall pathway signals detected. A comparison of the significant differently expressed genes from significant BioCarta, GO, KEGG and Reactome pathways converged on two genes *(PLAU* and *BIRC2* Fig 4B). We thus measured plasma UPA levels in the family members and observed that they were similar in cases and controls (Fig 2). Interestingly, *SERPINE1* was found at the intersection of significant BioCarta, GO and Reactome pathways (Fig 4B). Plasminogen activation potential assessed in plasma by the global fibrinolysis assay ECLT was significantly lower in cases than in controls (1200 +/-300 Min vs 458 +/-90 Min, p = 0.01) reinforcing the potential alteration the fibrinolysis cascade (Fig 2).

### Impact of MAST2R89Q mutation on the regulation of TFPI and SERPINE1 gene expression

Alterations in plasma levels of f-TFPI and PAI-1 measured in carriers of the MAST2R89Q mutation (Fig 2) associated with the regulatory role of MAST2 on the expression of the *TFPI* and *SERPINE1* genes (Fig 3C) led us to hypothesize an effect of the MAST2R89Q mutation on the expression of these genes. MAST2-dependent transcriptional regulation of *TFPI* and *SERPINE1* expression was studied in HEK293 cells expressing firefly luciferase under the control of *TFPI* promoter (TFPI-Luc) or destabilized EGFP under the control of *SERPINE1* promoter (PAI-1-dEGFP). Transfection of MAST2 specific siRNA, which moderately reduced MAST2 protein levels (Fig 5A), significantly increased *TFPI* promoter activity and reduced that of *SERPINE1* (Fig 5B), showing the ability of MAST2 to regulate the transcription of these genes. To investigate the effect of MAST2R89Q mutation on *TFPI* and *SERPINE1* transcription, wild type MAST2 and MAST2R89Q were overexpressed in HEK293 cells (Fig 5C) and their impact on the activity of *TFPI* and *SERPINE1* promoter was analyzed. *TFPI* promoter activity was lower and that of *SERPINE1* higher in cells overexpressing MAST2R89Q than in cells overexpressing wild type MAST2 (Fig 5D). This result suggests that the transcription of the *TFPI* and *SERPINE1* genes is altered in carriers of the MAST2R89Q mutation.

**Figure 5:**
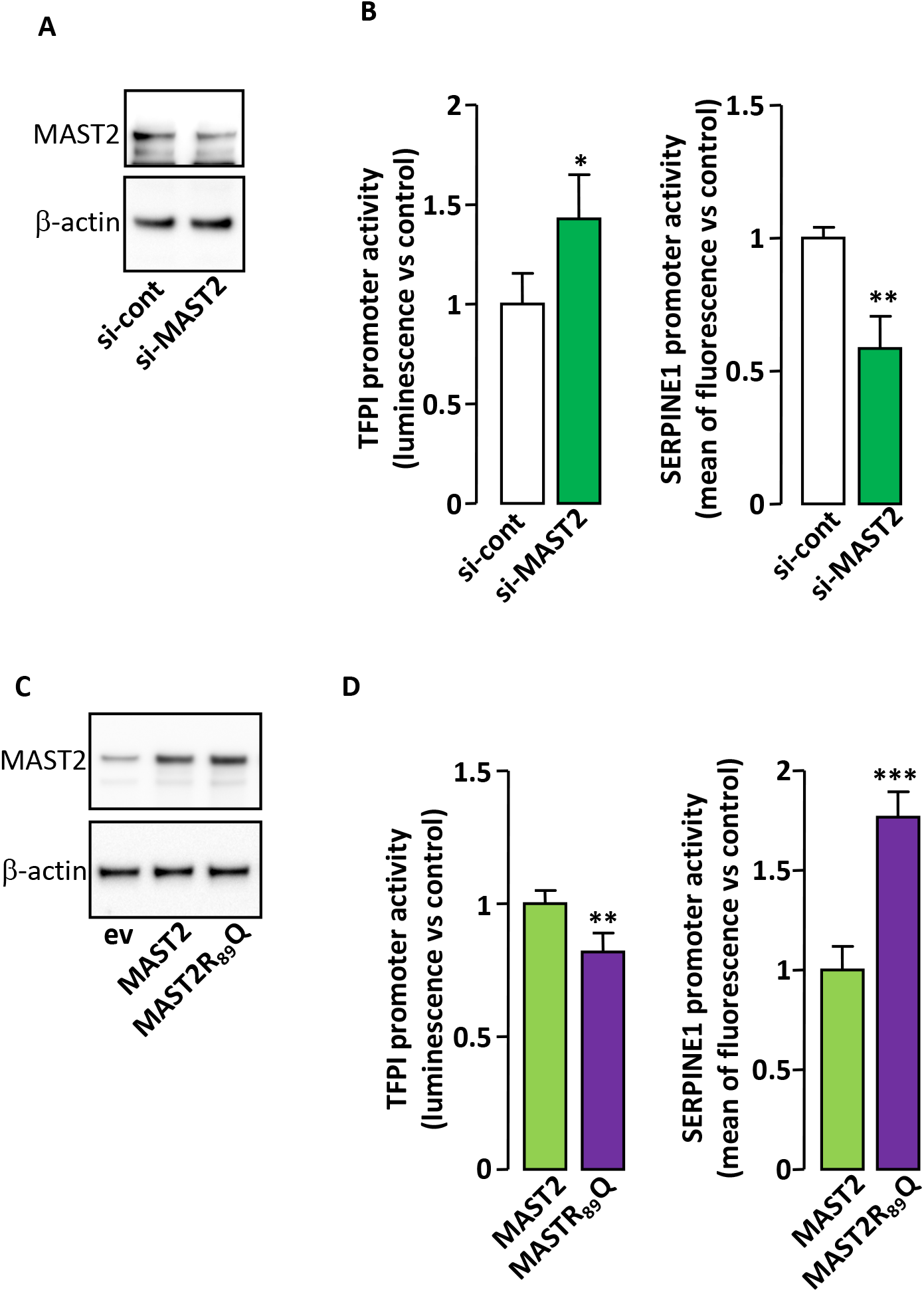
MAST2 regulates *TFPI* and *SERPINE1* promoter activity a). HEK293 cells were transfected with control siRNA or with the two MAST2 specific siRNA and MAST2 expression was analyzed by western blot 48 hours post transfection. The detection of β-actin attests for equal protein loading. b). HEK293 cells were cotransfected with control siRNA or with the two MAST2 specific siRNA together with TFPI (left) or PAI-1 (right) promoter reporter. 48 hours post transfection, the activity of the promoters was measured as described in material and methods section. Data are means ± standard deviation. c). HEK293 cells were transfected with empty vector (ev), wild type MAST2 or MAST2R89Q expression vector and MAST2 expression was analyzed by western blot 48 hours post transfection. The detection of β-actin attests for equal protein loading. d). HEK293 cells were cotransfected wild type MAST2 or MAST2R89Q expression vector together with TFPI (left) or SERPINE1 (right) promoter reporter then the activity of the promoters was measured as in figure 4b. Data are means ± standard deviation. Statistical analyses were made using Mann-Withney. **p<0.01; ***p<0.001: significant vs control.

## Discussion

Using a WES approach, we associate an extremely rare mutation to VTE in a thrombophilia family. The identification of this new mutation, R89Q located in exon2 of the *MAST2* gene, lead us to demonstrate for the first time that this gene is an important player of endothelium in VTE by regulating key coagulation and fibrinolysis parameters. The mutation is not present in whole genome/exome genomics resources neither in a collection of ~6,790 VTE cases and 5,970 controls specifically genotyped for it indicating that it is a very private mutation.

The identified mutation maps to the N-terminal domain of the MAST2 protein belonging to the Microtubule-associated serine/threonine-protein kinase (MAST) family. It is located in a highly conserved region across species and is predicted to be damaging by several prediction tools. The members of the MAST kinases family are characterized by the presence of a serine-threonine kinase domain, a second 3’ MAST domain with some similarity to kinase domains and a PDZ domain (9). They were demonstrated to link and phosphorylate the dystrophin/utrophin (DMD/UTRN) network with microtubule filaments via the syntrophins, modulating their affinities for associated proteins(10). This poorly understood family of proteins have been implicated in several human diseases, such as neurodegeneration and breast cancer (11). Our study is the first to report the involvement of MAST kinases in haemostasis. According to the Human Proteins Atlas (http://www.proteinatlas.org), MAST2 is ubiquitously expressed, mainly to the cytosol. By screening several coagulation parameters in the investigated family members, we observed that carriers of the mutation exhibited decreased TFPI plasma levels compared to non-carriers while the opposite pattern was observed for PAI-1. Those 2 molecules, which plasma levels are not affected by vitamin K antagonists, are both synthesized by endothelial cells and are major components of the coagulation/fibrinolysis process as TFPI is the primary inhibitor of the initiation of blood coagulation whereas PAI-1 is the main inhibitor of plasminogen activation. Microtubules are crucial components of the cytoskeleton which controls endothelial cell shape, migration and proliferation(12). In plasma, decreased in f-TFPI and increased in PAI-1 levels are both associated with the risk of thrombosis (13)(14)(15). According to these observations, we speculated that MAST2 might play an important role in regulating the hemostatic function of endothelial cells and that the identified variant interferes with the coagulation/fibrinolysis process affecting the risk of thrombosis. Knockdown experiments performed in ECV304 endothelial cell line followed by RNA-seq analysis confirmed that MAST2 participates to the regulatory mechanisms associated with the dissolution of fibrin clot through some mechanisms that need to be clarified. This is consistent with the altered ECLT in cases which is considered as a useful global fibrinolysis assay assessing plasminogen activation potential (16). Nevertheless, we showed that MAST2 acts as a negative regulator of *TFPI* expression and a positive regulator of that of *SERPINE1, PLAU* and *RHBLD2* coding for the transmembrane protease that cleaves THBD at the cell surface(17).

Interestingly, the lower activity of *TFPI* promoter in cells overexpressing MAST2R89Q than in those overexpressing wild type MAST2 suggests that the MAST2-dependent downregulation of *TFPI* transcription is increased by the mutation. The opposite effect was observed for *SERPINE1* promoter activity, suggesting that MAST2R89Q is more efficient in stimulating *SERPINE1* transcription than wild-type MAST2. Although *PLAU* expression is regulated by MAST2, the lack of significant differences in urokinase plasma levels between control individuals and patients with MAST2R89Q mutation suggests that it is not sensitive to the R89Q mutation. The MAST2-dependent regulation of *RHBDL2* expression and the impact this could have on THBD shedding is currently under investigation. However, sTHBD plasma levels are not different between control individuals and patients with MAST2R89Q mutation (data not shown), suggesting that in this particular context, modification of THBD cleavage by RHBDL2 is not sufficient to affect sTHBD plasma level.

Overall, these data reveal that MAST2 mutation is actively involved in the alterations of TFPI and PAI-1 expressions that are observed and would contribute to the pro-thrombotic state associated with this mutation.

The strength of the present study lies in the careful selection of the family studied. We selected relatives (across three generations) of a family with a very strong clinically prothrombotic phenotype and no detectable known inherited thrombophilia in order to increase our chance to identify new inherited thrombophilia that are likely to be rare. A special attention has been paid to identify, in this family, cases with a strong personal and certain (documented) history of VTE who are likely to have an underlying new inherited thrombophilia and controls with no coagulation abnormalities. As with any WES project, this strategy relies on the assumption that the variants to be identified reside in coding or untranslated regions, a realistic hypothesis, for at least some variants, since all thrombophilia anomalies identified so far are located in such regions. It is demonstrated that patients with unprovoked VTE at young age are more likely to have genetic risk factors of VTE. This family was also located in a geographically limited area, the Finistère department in Brittany: such families are rare, genetically poorly mixed with people outside of Brittany and more likely to share common genes from this geographical area.

Several limitations must be acknowledged: MAST2 mainly locates to the cytosol and it is then unclear how it could be directly involved in the regulation of *TFPI* and *SERPINE1* gene expressions. Moreover, out of the 15 subjects from the family studied, plasma samples were available in only 10 (3 cases and 7 controls). Further experimental studies are mandatory to decipher the underlying mechanisms. Another is about the lack of generalizability of the result. Indeed, we found no mutated patients in more than 6700 genotyped VTE patients suggesting that the mutation is private as observed with the FIX Padua (18). However, our results pave the way for adding *MAST2* to the list of genes to be sequenced and looked for in thrombophilia families with unprovoked VTE. More importantly, our study uncovers a new regulatory pathway of *TFPI* and *SERPINE1* expression by MAST2.

In conclusion, using a whole exome sequencing approach in a large pedigree affected with VTE, we identified an extremely rare mutation in the Microtubule-associated serine/threonine-protein kinase-2 *(MAST2)* gene, that is responsible for inherited thrombophilia through a not yet-fully characterized mechanism that involves modulation of the hemostatic balance of endothelial cells.

## Materials and Methods

### Recruitment of the family

Patient recruitment and her family members have been prospectively performed since 2010 in Finistère (France). All family members signed a written informed consent for genetic investigations. The recruitment had approval from the Brest Ethic Board. This family has been carefully selected from the index case with acute VTE and the following predefined criteria: a) VTE occurred at young age (< 50 years); b) was documented (i.e.; symptomatic proximal deep vein thrombosis and/or symptomatic pulmonary embolism objectively diagnosed according to validated criteria)(19)(20); c) was unprovoked (i.e.: VTE was not associated with clinical risk factors such as surgery, trauma or prolonged immobilization (≥ 72 hours) in the past 3 months, cancer in the past 2 years, chronic inflammatory illness, autoimmune disease, pregnancy, estrogen-containing pills, hormone replacement therapy); and d) with no established biological risk factors (antithrombin, protein C, protein S deficiencies, FV Leiden, prothrombin G20210A, lupus anticoagulant, anticardiolipin antibodies). This proband should also have at least two family members across three generations with a previous of symptomatic documented VTE with the same characteristics. All relatives (with or without history of VTE) have been invited to participate to the study excepted family members who were adopted or less than 16 years. All the participants have been asked to come to the Brest clinical center, at the CIC INSERM 1412, for clinical and biological screening of VTE. A predefined standardized form has been used to collect clinical data (demographic and clinical past history data, complete phenotype in the case of previous thrombosis, complete pedigree across three generations) and an extensive analysis of blood coagulation has been performed. Clinical characteristics of the subjects are reported in Table 2. A biobank including DNA, RNA, serum or citrate plasma samples stored at −80°C was set up for each family member.

**Table 2.** Clinical characteristics of the cases (i.e.; with VTE) in family members.

| Cases number | Gender | Relationship | WES | Age at VTE (years) | VTE type |
| --- | --- | --- | --- | --- | --- |
| 305000 | Female | Index case | Selected for WES | 34 | PE |
| 305001 | Female | Mother | Selected for WES | 27 | DVT |
|  |  |  |  | 45 | PE |
|  |  |  |  | 54 | PE |
| 305002 | Male | Maternal uncle | Selected for WES | 32 | DVT |
|  |  |  |  | 36 | PE + DVT |
| 305003 | Female | Maternal aunt | Selected for WES | 43 | PE + DVT |
| 305012 | Female | Maternal | Selected for replication | 35 | PE |

### Measurement of coagulation and fibrinolytic traits

Several coagulation and fibrinolytic parameters, not belonging to the classical thrombophilia screening, were measured in 10 family members with plasma available (3 cases and 7 controls). The 3 cases were on warfarin whereas none of the controls were on anticoagulant treatment at the time of the sampling.

Activated partial thrombin time (aPTT) and prothrombin time (PT), fibrinogen, factor II, factor V and factor X were measured in plasma with automated coagulometers (STA-Revolution, Diagnostica Stago, Asnières, France) using established commercial assays. Free tissue factor pathway inhibitor (f-TFPI) plasma levels were measured with the Asserachrom Free TFPI enzyme immunoassay from Diagnostica Stago (Asnieres, France) and plasma PAI-1 activity with the Zymutest PAI-1 activity from Hyphen Biomed (Neuville-sur-Oise, France). Urokinase (uPA) plasma levels were measured with the uPA ELISA kit (Abcam, Cambrige, UK). The euglobulin clot lysis time (ECLT) assay has been performed as previously described (21).

### DNA analysis

Fifteen family members were studied (see genealogical tree in Fig 1). Relatives with a previous VTE event as defined by our inclusion criteria were considered as cases. From 5 cases with DNA available, 4 were selected for being whole exome sequenced. These were the most distant VTE relatives. From the remaining family members who have not had any previous episode of venous or arterial thrombosis and that presented with normal thrombin generation test and a normal leg doppler ultrasound (individuals considered as controls), we selected for WES two that were expected to be as genetically close as possible from cases (Fig 1). Clinical characteristics of the WES participant are shown in Table 2.

A 2 × 100 paired-end sequencing was performed on an Illumina HiSeq2000 instrument at the IGBMC platform (http://www.igbmc.fr/).

### Bioinformatic WES workflow

FastQ sequences were aligned to the hs37d5 version of the human reference genome hg37 with BWA-MEM algorithm (22) of the Burrows-Wheeler Aligner.

All duplicated reads were marked using picard-tools-v1.119 (http://broadinstitute.github.io/picard/) and sorted using samtools-v1.3.1(23). Base quality check was performed prior to the variant calling with GATK - BaseRecalibrator (GenomeAnalysisTK-v3.3-0).

Single nucleotide variants (SNVs) and small insertions/deletions (INDELs) were called following GATK’s Best Practices (https://software.broadinstitute.org/gatk/best-practices) with HaplotypeCaller. After the GATK’s VQSR step, «PASS» variants were annotated using ANNOVAR(24) package.

As a strategy to identify the culprit variant, we first prioritized variants that were likely functional (stop loss/stop gain, frameshift insertion/deletion, non-synonymous and splicing variants) and that were carried by all VTE cases and not by any of the two controls. Second, a bioinformatics search in public genomic data repositories (eg dbSNP, NCBI, Ensembl, 1000 genomes, ExAC (http://exac.broadinstitute.org) and FrEx (http://lysine.univ-brest.fr/FrExAC/) was carried out on prioritized variants to select as suggestive candidates those that have not been reported, or reported to be at low frequencies (<1‰). We also took into account the predicted deleteriousness of selected candidates using *in silico* tools such as SIFT, PolyPhen and CADD-v1.2 (25) to further reduce the number of candidates.

Candidate variants identified from this multi-step strategy were then genotyped in all family members for whom DNA was available, including an additional set of 1 case and 8 controls beyond the initial group of individuals that were part of the WES experiment.

### Validation of the impact of the newly identified variants in other cohorts of patients with VTE

Identified candidate variants were genotyped in additional samples of VTE patients and healthy individuals in order to get a more accurate estimation of their allele frequencies and more evidence for the putative one. Several French studies for VTE were investigated. These were:

- the “FIT study”, a prospective trans-sectional cohort assessing the risk of VTE in 2617 first-degree relatives of 507 index cases with VTE (507 families). Description of the cohort has been previously reported (26).
- the MARTHA, FARIVE and EDITH case-control studies for VTE, totaling 4,173 VTE cases and 5,970 healthy individuals whose detailed description has already been reported elsewhere (5).

We retrieved information about variant rs1387081220 from large scale genotyping projects which, in aggregate, comprised information from 345,939 genetically independent individuals, with 60.26%, 12.51% and 12.35% being of Caucasoid, Asian and African ethnicity groups respectively. A single copy of the minor allele of rs1387081220 was observed in two female individuals from the gnomAD dataset (one Caucasian and one African) but was unobserved in the remaining 345,937 individuals across all collections. However, no phenotype information can be obtained from the gnomAD participants. The results of the look-up of genotypes is summarised in S2A-B Table.

### Functional characterization of the newly identified genetic variants

The role of MAST2 in endothelial function was assessed by small interference RNA (siRNA) silencing. Comparison of the endothelial transcriptome of ECV304 cells cotransfected with two MAST2 siRNA leading to a significant downregulation of MAST2 expression (around 80 % reduction of mRNA levels) compared to ECV304 cells transfected with control siRNA was performed using RNA-Seq.

Total RNA extracted from transfected ECV304 cells with control or Mast2 siRNA were used to prepare libraries using the NEBNex Ultra II Direction RNA library prep kit for Illumina protocol according to supplied recommendations. Libraries were sequenced on an Illumina NovaSeq instrument using a paired end sequencing of 2 x 100bp. Sequencing and bioinformatics analysis of sequenced data were outsourced at the Integragen company (https://www.integragen.com/). Base calling was performed using Illumina Real Time Analysis (3.4.4) with default parameters. Fastq files were aligned to the reference Human genome hg38 with STAR (27) with the following parameters: --twopassMode Basic -- outReadsUnmapped None --chimSegmentMin 12 --chimJunctionOverhangMin 12 - alignSJDBoverhangMin 10 --alignMatesGapMax 200000 --alignIntronMax 200000 - chimSegmentReadGapMax parameter 3 --alignSJstitchMismatchNmax 5 -1 5 5 --quantMode GeneCounts --outWigType wiggle --sj dbGTFtagExonParentGene gene_name). Reads mapping to multiple locations were removed. Gene expression were quantified using the full Gencode v31 annotation. STAR was also used to obtain the number of reads associated to each gene in the Gencode v31 database (restricted to protein-coding genes, antisense and lincRNAs). The Bioconductor *DESeq* package (28) was used to import raw counts for each sample into R statistical software and extract the corresponding count matrix. After normalizing for library size, the count matrix was normalized by the coding length of genes to compute FPKM scores (number of fragments per kilobase of exon model and millions of mapped reads).

#### Plasmid constructs

Human TFPI promoter Firefly-luciferase reporter (TFPI-Luc), a pGL3-Basic vector containing a fragment of TFPI promoter from −1224 to +45, was previously described (29). PAI-1 promoter (−483/+75) (30) was PCR amplified from THP1 genomic DNA then inserted in place of the CMV promoter in the pd4EGFP plasmid (Clontech) that contains the sequence coding for a destabilized version of EGFP. The resulting vector was named PAI-1-dEGFP. GFP-tagged MAST2 expression vector (RG206492 form Origene) was used as template to PCR amplify only the full-length coding sequence of MAST2 (without the GFP coding sequence), which was then cloned into pCDNA3 vector using the In-Fusion PCR cloning system (Clontech).

#### Cell culture and transfection

HEK293 cells (Griptite 293 MSR) were from Thermo Fisher Scientific. ECV304 endothelial (31) cells were from ATCC. The cells were maintained in culture as described by the manufacturers. Transfections of plasmid DNA were performed with PolyJet reagent (SignaGen Laboratories, Rockville, MD, USA). Transfections of siRNA alone or in association with plasmid DNA were performed with JetPRIME reagent (Polyplus transfection), as specified by the manufacturers. MAST2 specific siRNA (#4392420; ID: s42 and # AM51331; ID: 975) and negative control siRNA were from Thermofisher.

#### Western Blot

Identical amounts of total protein were heat-denatured and reduced (70°C; 10min) then submitted to SDS-PAGE separation on 4-12 % gradient NuPAGE gels (Life Technologies, Saint Aubin, France) and transferred to polyvinylidene fluoride membranes. The membranes were blocked for 1 hour in 5 % BSA solution, immunodetections were performed with MAST2 (ab209079 from Abcam) and β-actin (13E5 Cell Signaling Technology) specific antibodies. Image acquisition was performed by using a chemiluminescent CCD imager Image Quant LAS 4000 (GE Healthcare, Velizy-Villacoublay, France).

#### Gene reporter experiments

Promoter activities were measured 48 hours after cell transfection. For the study of PAI-1 promoter activity assay, HEK293 cells transfected with human PAI-1-dEGFP were detached by incubation in PBS without calcium and magnesium and EGFP fluorescence was measured by flow cytometry (Accuri C5, BD Biosciences). PAI-1 promoter activity was calculated as the means of fluorescence corrected for the value obtained with cells transfected with an empty vector and expressed as fold change compared to the control situation. For the study of TFPI promoter activity assay, HEK293 cells were cotransfected with human TFPI-Luc and SV40-driven Renilla luciferase coding vectors. Firefly and Renilla luciferase were measured in cell lysates using a luminometer (EnSight Multimode plate reader, Perkin Elmer). TFPI promoter activity was calculated as the ratio Firefly/Renilla luciferase and expressed as fold change compared to control.

#### Real-Time PCR analysis

Total RNA was extracted using the Nucleospin RNA Kit (Macherey-Nagel, Hoerdt, France), cDNA was synthesized from 0.5 μg of RNA using MMLV reverse transcriptase (Life Technologies, Saint Aubin, France) and used for PCR amplification. RT-PCR were performed on the LightCycler 480 instrument (Roche Applied Science, Meylan, France) using the Eva Green MasterMix (Euromedex, Souffelweyersheim, France). The comparative Ct method (2^-(ΔΔCT)^) was used to calculate the relative differences in mRNA expression. The acidic ribosomal phosphoprotein P0 was used as housekeeping gene. Primers sequences are available upon request. Changes *were* normalized to the mean of *control values*, which *were set* to 1.

#### Statistical Analyses

All data but RNA sequencing data were analyzed with GraphPad Prism software and individual *statistical two-sided tests used* are identified in the *figure legends*. P values ≤ 0.05 were considered statistically significant.

Differential analysis of RNA Seq data was performed using the Bioconductor *limma* package (32) on voom-transformed data in order to identify gene expression levels impacted by *MAST2* siRNA silencing. A False Discovery Rate (FDR) threshold of 1% was used to identify candidate mRNAs associated with *MAST2* knockdown. A functional enrichment analysis using the Database for Annotation, Visualization and Integrated Discovery software (DAVID;(8)), was performed on the most dysregulated genes to get insight into the biological pathways perturbated by *MAST2* knockdown. For this analysis, the GO, KEGG, REACTOME, and PANTHER databases were interrogated.

## Data Availability

All data in the manuscript are available upon request to the corresponding author.

## Acknowledgements

GV.S was financially supported by the GENMED Laboratory of Excellence on Medical Genomics [ANR-10-LABX-0013]. Sequencing was funded by the Fondation pour les Maladies Rares and was performed by the IGBMC Microarray and Sequencing platform, a member of the “France Génomique” consortium (ANR-10-INBS-0009). DA.T was financially supported by the «EPIDEMIOM-VTE» Senior Chair from the Initiative of Excellence of the University of Bordeaux.

The NIHR BioResource-Rare Diseases projects were approved by Research Ethics Committees in the UK and appropriate national ethics authorities in non-UK enrolment centers. This research was made possible through access to the data and findings generated by the 100,000 Genomes Project. The 100,000 Genomes Project is managed by Genomics England Limited (a wholly owned company of the Department of Health and Social Care). The 100,000 Genomes Project is funded by the National Institute for Health Research and NHS England. The Wellcome Trust, Cancer Research UK and the Medical Research Council have also funded research infrastructure. The 100,000 Genomes Project uses data provided by patients and collected by the National Health Service as part of their care and support.

We thank the Professor Sandset for the kind gift of the PGL3-TFPI promoter vector.

## Supporting information

**S1 Table. Genotype distribution of the *MAST 2* variant in family members**.

**S2 Table. Reference genotyping datasets and genetically inferred ethnicity**.

**S3 Table. Full list of differential association p-value of RNA-Seq based analysis to compare the mRNA expression profiles of ECV 304 endothelial cells, with those of *MAST2* knockdown cells**.

**S4 Table. Pathway analysis applied to the top 100 most significantly differentiated mRNA expressions according to *MAST2* silencing**.

## Notes

### Competing Interest Statement

The authors have declared no competing interest.

### Clinical Trial

The study was not registered as it is not a clinical trial

### Author Declarations

All family members signed a written informed consent for genetic investigations. The recruitment had approval from the Brest Ethic Board.

